# Biomedical knowledge gaps, supernatural causal beliefs and the traditional-first treatment pathway sustaining onchocerciasis transmission despite mass drug administration in Bo District, Sierra Leone: an explanatory sequential mixed-methods study

**DOI:** 10.64898/2026.06.18.26355997

**Authors:** Augustine Bob Moseray, Alhaji Brima Gogra, Rashid Ansumana

**Affiliations:** Department of Public Health, School of Community Health Sciences, Njala University, Bo Campus, Bo, Sierra Leone; Ernest Bai Koroma University of Science and Technology, Port Loko Campus, Port Loko, Sierra Leone; School of Medical Health Sciences, Njala University, Bo Campus, Bo, Sierra Leone

## Abstract

Onchocerciasis (river blindness) persists in Bo District, Sierra Leone, despite two decades of ivermectin-based mass drug administration (MDA). Whether community knowledge, causal beliefs and treatment-seeking continue to favour transmission has not been quantified at scale. Using an explanatory sequential mixed-methods design, we surveyed 1,500 adults across five endemic chiefdoms (Bagbwe, Baoma, Gbo, Selenga, Tikonko) using multistage cluster sampling, and conducted 11 key informant interviews and 8 focus group discussions analysed thematically. Biomedical perception was measured with closed items summed into a composite score (range 0–3); the principal behavioural outcome was traditional-first treatment (use of traditional remedies before modern care). Quantitative data were analysed with chi-square tests, logistic regression, and ROC analysis. Biomedical understanding was moderate and incomplete: only 46.5% correctly identified the blackfly vector and 38.1% judged community awareness sufficient, while supernatural or moral attributions accounted for 68.1% of social explanations and half the sample (50.3%) endorsed a traditional or spiritual origin (mean composite perception 1.77/3). Perception did not vary across chiefdoms (all Cramér’s V < .06); the only consistent correlate was prior experience of the disease, which raised the odds of adequate perception (OR = 0.77 for inadequate perception, p = .016). Traditional-first treatment was the modal pathway (55.3%), care-seeking was reactive (70.1% acted only on symptom onset) and socially governed (71.3% consulted family or peers), and traditional healers were present in most communities (56.9%). A flagship logistic model predicting traditional-first treatment had negligible explanatory power (McFadden R² = .005; likelihood-ratio p = .737; AUC = 0.544), with only distrust of modern medicine showing a directional effect (OR = 1.33, p = .065). Qualitative accounts explained these patterns as a coherent cultural logic rather than ignorance, rooted in naming the disease for water, spiritual aetiologies, the invisibility of the parasite, and the complementary role of healers. Aetiology-aligned health communication, engagement of traditional healers and gender-sensitive MDA delivery are needed to close the residual transmission gap.

**Author summary:** River blindness is a parasitic disease spread by blackflies that breed in fast-flowing rivers. For twenty years, communities in Bo District, Sierra Leone, have received a yearly preventive medicine (ivermectin), yet the disease has not gone away. We wanted to understand what people in these communities know and believe about the disease, and where they go first when they fall ill. We asked 1,500 adults in five rural areas and held in-depth interviews and group discussions. Fewer than half knew the blackfly spreads the disease, and most explained it through curses, witchcraft or the spirit of the water rather than a parasite they cannot see. More than half tried traditional remedies before going to a clinic. Importantly, this choice was not predicted by a person’s sex, age, education or village it was a shared cultural default driven by belief and by distrust of modern services. People did not reject medicine; they combined it with healing that addresses the spiritual meaning of illness. Elimination programmes are therefore unlikely to succeed by distributing drugs alone. They must speak to local explanations of the disease, work with traditional healers, and remove the practical and gender-related barriers that keep modern treatment out of reach.

## Introduction

Onchocerciasis, or river blindness, is a chronic filarial disease caused by *Onchocerca volvulus* and transmitted by *Simulium* blackflies that breed in fast-flowing rivers (Basáñez et al., 2006; Frallonardo et al., 2022). The disease produces intense itching, disfiguring skin disease, visual impairment and blindness, and is associated with epilepsy and excess mortality in heavily infected populations (Basáñez et al., 2006; Schmidt et al., 2022). Sub-Saharan Africa carries almost the entire global burden, and Sierra Leone has historically been among the most intensely affected countries, with the riverine ecology of districts such as Bo sustaining vigorous vector breeding (Post & Crosskey, 1985; Schmidt et al., 2022; Zouré et al., 2014).

The principal control strategy is community-directed treatment with ivermectin (CDTI), delivered through annual or biannual mass drug administration (MDA) (Koroma et al., 2018). Two decades of MDA have markedly reduced infection intensity in Sierra Leone, yet recent serological and entomological assessments indicate that transmission has not been interrupted in several endemic foci, and the country remains short of the 2030 elimination target (Gebrezgabiher et al., 2019; Kargbo-Labour et al., 2024; Koroma et al., 2018). As programmes shift from morbidity control to transmission elimination, the residual drivers of persistence suboptimal therapeutic coverage, systematic non-participation, and the social and cognitive determinants of treatment-seeking have become the decisive obstacles (Colebunders et al., 2018; Gebrezgabiher et al., 2019).

A substantial literature shows that biomedical knowledge of onchocerciasis in endemic communities is frequently partial: people recognise the disease and its symptoms but misattribute its cause, commonly to supernatural, moral or non-parasitic natural forces [10,11,12](Okyere et al., 2024). Where a disease is understood as a curse or a spiritual affliction, the perceived benefit of a biomedical drug is diminished and recourse to traditional or spiritual healing is a rational choice within that explanatory frame [12,13]. The result is a pluralistic therapeutic landscape in which biomedical, traditional and spiritual options coexist and are used concurrently or sequentially, often with traditional care sought first [14,15]. These socio-behavioural determinants causal beliefs, trust in distributors, and gendered access barriers have been repeatedly identified as programmatic vulnerabilities, yet they remain under-investigated in Sierra Leone, and almost never measured at a scale that permits robust modelling [9,16](Colebunders et al., 2018).

This study addresses that gap. Using an explanatory sequential mixed-methods design among 1,500 adults across five endemic chiefdoms of Bo District, we (i) quantify biomedical perception and causal beliefs about onchocerciasis; (ii) test whether perception varies by chiefdom and socio-demographic group, and identify its determinants; (iii) characterise healthcare-seeking behaviour and the prevalence of the traditional-first treatment pathway; (iv) model the predictors of traditional-first treatment; and (v) use qualitative accounts to explain the observed patterns. The analysis is organised by an integrated Health Belief Model–Socio-Ecological Model (HBM–SEM) framework that situates individual cognition within community and structural context [16,17].

## Methods

### Ethics statement

Ethical approval was granted by the Njala University Postgraduate Review Board, with additional permissions from the Bo District Health Management Team and the relevant chiefdom authorities. All participants provided voluntary informed consent. For non-literate participants, consent was documented by thumbprint witnessed by an independent community member, an oral-consent procedure approved by the review board. No individually identifying information is presented.

### Study design

An explanatory sequential mixed-methods design was used: a quantitative cross-sectional survey was conducted and analysed first, followed by a qualitative phase that explained and elaborated the quantitative findings [18,19]. The design is grounded in a pragmatic paradigm that prioritises the research question over methodological allegiance [18].

### Study area and population

The study was conducted in Bo District, Southern Province, Sierra Leone. The district has a tropical monsoon climate and an extensive hydrological network, including the Sewa River, that creates abundant *Simulium damnosum* breeding habitat and underpins onchocerciasis endemicity [4,5]. The economy is predominantly agrarian and riverine. Twenty-six rural villages across five endemic chiefdoms (Bagbwe, Baoma, Gbo, Selenga and Tikonko) were selected in consultation with the Bo District Health Management Team on the basis of documented endemicity, MDA history, coverage challenges and ecological diversity. The survey population comprised all permanently resident adults aged 18 years and older.

### Sample size and sampling

Sample size was based on a single-proportion formula (p = 0.5, d = 0.05, 95% confidence), adjusted for a design effect of 1.5 and 10% non-response [20,21]. To permit adequately powered chiefdom-level disaggregation (≥300 respondents per chiefdom to detect a 15-percentage-point difference at 80% power), the target was set at 1,500. A multistage cluster design was used: chiefdoms were purposively selected (stage 1); villages were selected stratified by ecological and socio-economic characteristics (stage 2); households were sampled systematically (stage 3); and one eligible adult per household was selected using the Kish grid (stage 4).

### Measures

Biomedical perception was captured with closed items covering knowledge of the blackfly vector, ability to describe symptoms, recognition of environmental factors, and judged sufficiency of community awareness; correct biomedical responses were summed into a composite perception score (range 0–3). Causal beliefs were measured by items on the dominant social attribution applied to affected persons, the perceived primary cause, and endorsement of a traditional or spiritual origin. Healthcare-seeking was operationalised principally as traditional-first treatment (use of traditional remedies before modern care; yes/no), alongside items on care-seeking triggers, social influences on care, the presence of traditional healers, perceived convenience of facilities, perceived urban–rural disparity, and barriers to modern treatment. All indicator items were closed, single-response questions answered by every respondent. Composite scores are formative sums of their constituent items rather than latent psychometric scales.

### Quantitative analysis

Data were analysed in JASP (v0.18). Associations between categorical variables were tested with the Pearson chi-square test, using Cramér’s V (or φ for 2×2 tables) as the effect-size measure. Differences in the composite perception score across chiefdoms were tested by one-way ANOVA. Binary logistic regression was used to identify predictors of (i) adequate biomedical perception and (ii) traditional-first treatment, with model fit and discrimination assessed by the likelihood-ratio test, McFadden and Nagelkerke pseudo-R², classification accuracy, sensitivity, specificity, and the area under the receiver-operating-characteristic curve (AUC). Significance was set at p < 0.05.

### Qualitative phase

Eleven key informant interviews (community drug distributors, community health officers, village chiefs, a traditional healer and the district NTD focal person) and eight focus group discussions were conducted in Mende and Krio, audio-recorded, transcribed and translated. Transcripts were analysed thematically in MAXQDA 2022 using a combined inductive deductive approach, with deductive codes derived from the HBM–SEM framework. Reflexivity was maintained through a systematic journal and deliberate epistemic humility toward spiritual explanations, which were treated as data to be understood rather than misconceptions to be corrected. Quantitative and qualitative findings were integrated through joint-display tables aligning each theme with the corresponding statistical result and framework construct.

## Results

### Sample characteristics

All 1,500 sampled adults participated. The sample was balanced by sex (51.0% female, 49.0% male) and predominantly young (54.9% aged 18–37). Farming was the leading occupation (51.7%), followed by fishing (17.6%). Educational attainment was low: 27.9% had no formal education and only 11.7% had tertiary education. The population was 56.6% Muslim and 43.4% Christian, with a mean household size of 6.9.

### Biomedical knowledge and causal beliefs

Biomedical understanding was moderate and incomplete (Fig 1; Table 1). Fewer than half of respondents (46.5%) correctly identified the blackfly as the vector, and only 38.1% judged community awareness to be sufficient, although 63.5% could describe the symptoms and 71.7% recognised environmental factors. Causal beliefs leaned heavily on non-biomedical frameworks: supernatural and moral attributions (curse, witchcraft, moral failing) accounted for 68.1% of the social explanations applied to affected persons, and half the sample (50.3%) endorsed a traditional or spiritual origin for the disease. The composite biomedical-perception score averaged only 1.77 of a possible 3 (SD 0.86), confirming that partial, symptom-anchored knowledge coexists with strong non-biomedical aetiologies.

**Fig 1.**
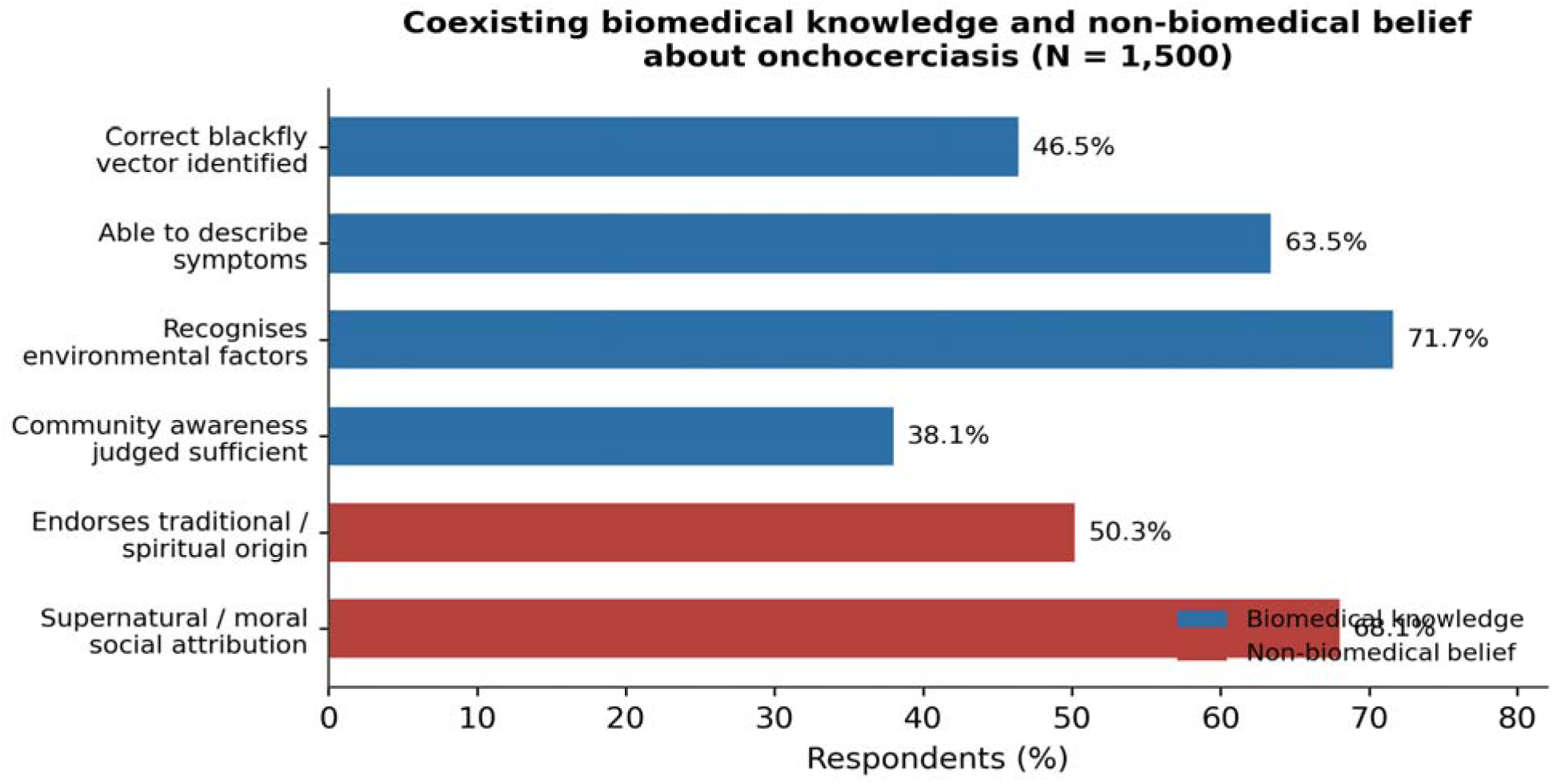
Biomedical knowledge and causal beliefs about onchocerciasis among 1,500 adults in five endemic chiefdoms of Bo District. Bars show the proportion endorsing each knowledge or belief indicator; biomedical-knowledge items and non-biomedical causal beliefs are distinguished by colour.

**Table 1.** Biomedical perception indicators, causal beliefs and composite perception score (N = 1,500).

| Indicator | n | % |
| --- | --- | --- |
| Correctly identifies blackfly vector | 698 | 46.5 |
| Able to describe symptoms | 953 | 63.5 |
| Recognises environmental factors | 1,076 | 71.7 |
| Judges community awareness sufficient | 572 | 38.1 |
| Endorses traditional/spiritual origin | 755 | 50.3 |
| Supernatural/moral social attribution | 1,021 | 68.1 |

*Composite biomedical-perception score (0–3): mean 1.77 (SD 0.86). The composite is a formative sum of the biomedical-knowledge items.*

### Determinants of biomedical perception

Perception was distributed evenly across the district: it did not vary significantly across the five chiefdoms, and every chiefdom-level association was trivial (all Cramér’s V < .06). The one consistent individual-level signal was prior experience of the disease. Respondents who had previously encountered a case held more accurate transmission views (χ²(3) = 10.25, p = .017, V = .083) and were less likely to express the dominant supernatural social attribution (χ²(3) = 9.13, p = .028, V = .078). In a logistic model for inadequate perception (Table 2), prior case exposure was protective (OR = 0.77, p = .016), while male sex (OR = 1.25, p = .037) and secondary education (OR = 1.33, p = .046) were associated with greater odds of inadequate perception modest effects against a backdrop of population-wide partial knowledge.

**Table 2.** Associations and logistic predictors of biomedical perception (N = 1,500).

| Predictor / test | Statistic | p |
| --- | --- | --- |
| Perception × chiefdom (all items) | Cramér's $V < .06$ | ns |
| Transmission view × prior case exposure | $\chi^2(3) = 10.25$ | .017 |
| Social attribution × prior case exposure | $\chi^2(3) = 9.13$ | .028 |
| Prior case exposure (logistic; inadequate perception) | OR = 0.77 | .016 |
| Male sex (logistic; inadequate perception) | OR = 1.25 | .037 |
| Secondary education (logistic; inadequate perception) | OR = 1.33 | .046 |

*ns = not significant. Outcome for the logistic model is inadequate biomedical perception (composite score below the sample median); odds ratios > 1 indicate higher odds of inadequate perception.*

### Healthcare-seeking and the traditional-first pathway

Care-seeking was reactive and socially governed (Fig 2; Table 3). Symptom onset, rather than prevention, was the trigger for 70.1% of respondents, and 71.3% sought counsel from family or peers before deciding on care; peer influence was acknowledged by 61.5%. Traditional-first treatment trying traditional remedies before modern care was the modal pathway, reported by 55.3% of respondents, and traditional healers were present and active in most communities (56.9%). The formal sector was perceived as poorly accessible: only 47.4% found facilities convenient and 63.0% perceived an urban–rural access disparity. The leading barriers to modern treatment were facility scarcity (24.1%), financial constraint (20.3%), transport (19.9%), geographic distance (19.1%) and stigma (16.7%).

**Fig 2.**
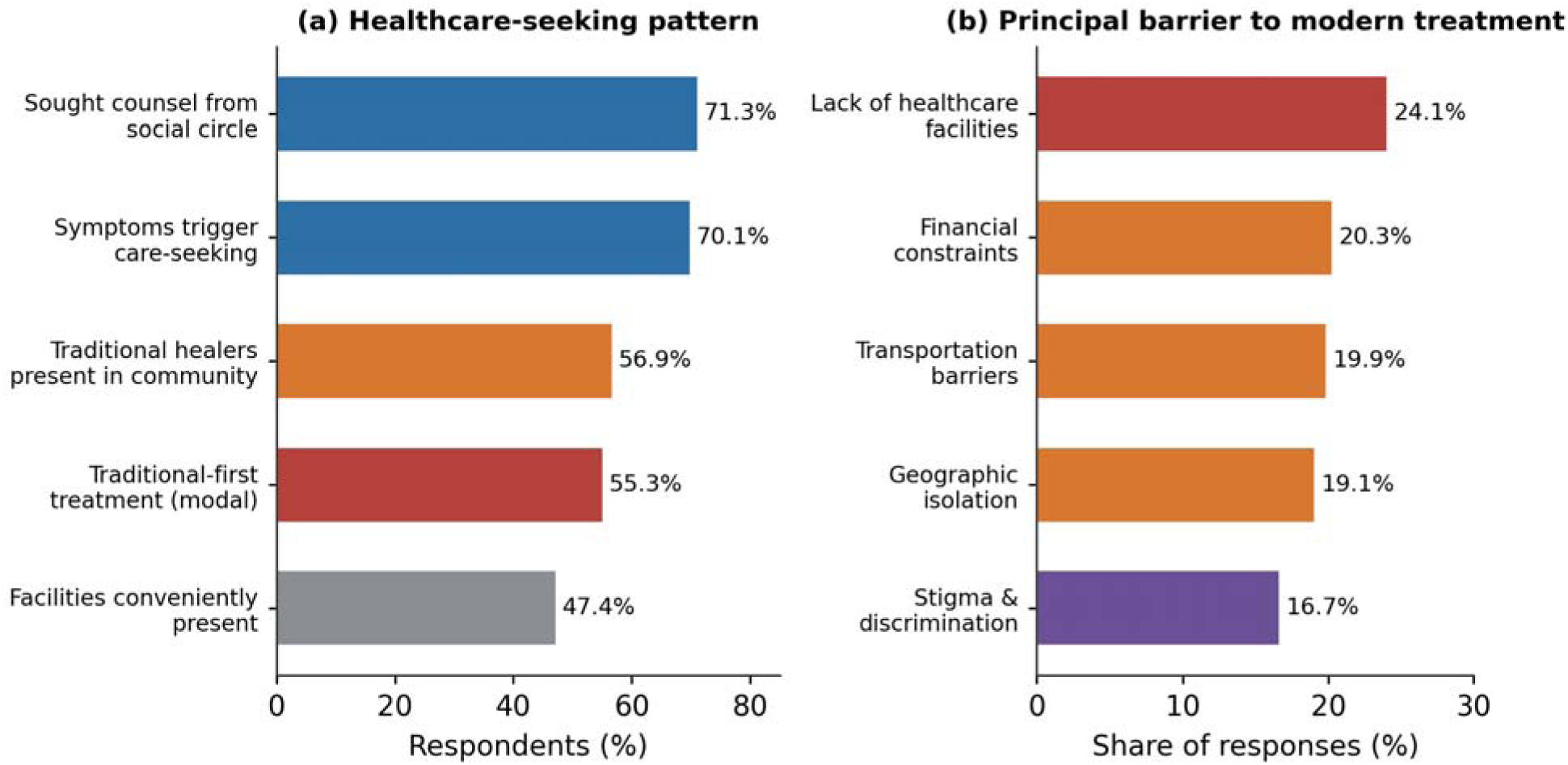
Healthcare-seeking behaviour and barriers to modern treatment for onchocerciasis (N = 1,500). The traditional-first pathway and the structural and social factors shaping care-seeking are shown alongside the principal reported barriers to modern-treatment uptake.

**Table 3.**
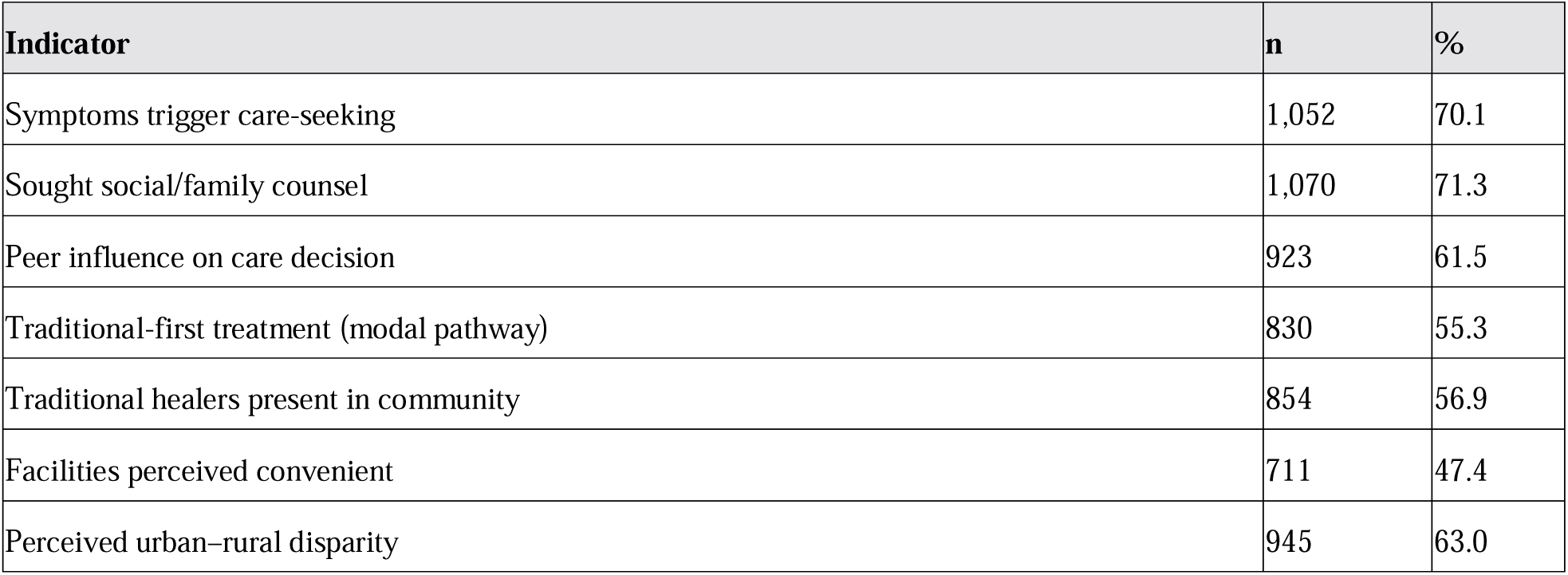
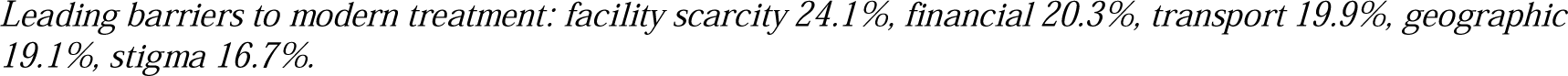
Healthcare-seeking indicators and barriers to modern treatment (N = 1,500).

| Indicator | n | % |
| --- | --- | --- |
| Symptoms trigger care-seeking | 1,052 | 70.1 |
| Sought social/family counsel | 1,070 | 71.3 |
| Peer influence on care decision | 923 | 61.5 |
| Traditional-first treatment (modal pathway) | 830 | 55.3 |
| Traditional healers present in community | 854 | 56.9 |
| Facilities perceived convenient | 711 | 47.4 |
| Perceived urban–rural disparity | 945 | 63.0 |

Leading barriers to modern treatment: facility scarcity 24.1%, financial 20.3%, transport 19.9%, geographic 19.1%, stigma 16.7%.

### Predictors of traditional-first treatment: the flagship model

No sociodemographic variable chiefdom, sex, education or religion was significantly associated with traditional-first treatment, and all effect sizes were trivial (V ≤ .036). A multivariable logistic regression (Fig 3; Table 4) confirmed that treatment choice was largely unpredictable from the measured variables: the model was not significant overall (likelihood-ratio p = .737) and explained almost none of the variance (McFadden R² = .005). Discrimination was poor (AUC = 0.544), and although overall classification accuracy was 55.3%, this reflected the model defaulting to the majority class sensitivity for the traditional-first pathway was 90.0% but specificity was only 12.4%. The single predictor approaching significance was distrust of modern medicine, which raised the odds of traditional-first treatment by about a third (OR = 1.33, 95% CI 0.98–1.81, p = .065); preference for traditional healers showed a similar but weaker trend. The substantive message is that the traditional-first pathway is a shared cultural default driven, to the limited extent it is predictable, by belief and trust rather than by social position most of what determines treatment choice lies in the structural and cultural environment rather than in individual attributes.

**Fig 3.**
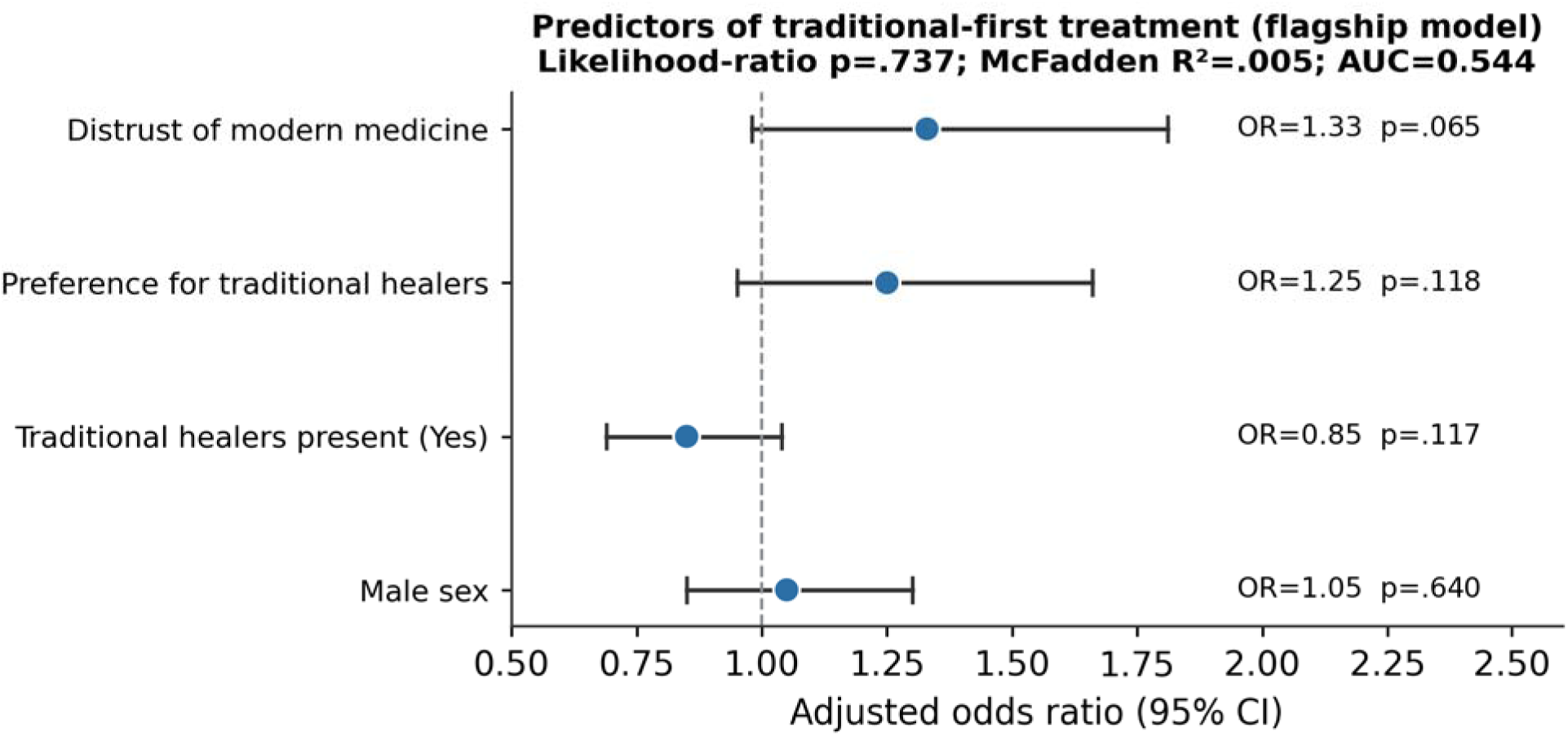
Adjusted odds ratios for predictors of traditional-first treatment (flagship logistic model; N = 1,500). Points are odds ratios and horizontal lines 95% confidence intervals; the vertical reference line marks OR = 1. No predictor reached statistical significance.

**Table 4.** Flagship logistic regression predicting traditional-first treatment and model performance (N = 1,500).

| Predictor | OR | 95% CI | p |
| --- | --- | --- | --- |
| Distrust of modern medicine | 1.33 | 0.98–1.81 | .065 |
| Preference for traditional healers | 1.25 | 0.95–1.66 | .118 |
| Traditional healers present | 0.85 | 0.69–1.04 | .117 |
| Male sex | 1.05 | — | .640 |

*Model fit: likelihood-ratio $p = .737$ ; McFadden $R^2 = .005$ . Discrimination: AUC = 0.544; accuracy 55.3%; sensitivity 90.0%; specificity 12.4%.*

### Qualitative explanation of the perception–treatment patterns

The qualitative phase explained the quantitative patterns as a coherent cultural logic rather than simple ignorance. Three knowledge themes accounted for the partial-perception finding. Experiential knowledge without biomedical understanding (Theme 1) was captured in the local name for the disease, *krijin weh de na wata* (“the sickness that comes from the water”), which correctly associates the illness with riverine environments but attributes cause to the water or its spirits rather than the vector: one participant noted that the blackfly bites, but that the real illness comes from the spirit of water (Male, FGD, Selenga). Spiritual and cosmological frameworks (Theme 2) were invoked for severe or disfiguring presentations, with depigmentation read as a curse that, as one woman put it, no hospital medicine can lift (Female, FGD, Bagbwe). The invisibility of the parasite (Theme 3) was an explicit ground for scepticism toward the biomedical account, voiced by a traditional healer who asked how a tiny fly could place a worm under the skin he had never seen (KII, Tikonko).

Treatment behaviour was explained by three further themes. Non-linear, pluralistic pathways (Theme 8) were the norm: participants described using herbs first, then the clinic when itching persisted, while also praying, treating biomedical and traditional care as complementary rather than mutually exclusive (Male, FGD, Tikonko). Trust deficits in service quality (Theme 10) undermined the formal sector, with distributors perceived as giving the drug without explanation, prompting doubts about their competence (Male, FGD, Tikonko). Gender-specific structural barriers (Theme 11) meant standard morning distribution coincided with women’s water-collection and food-preparation duties, so that distributors had often left before women returned (Female, FGD, Baoma), an exclusion compounded by inconsistent messaging about ivermectin in pregnancy. Finally, the complementary role of healers (Theme 12) was articulated by a healer who distinguished the hospital’s treatment of the body from his own treatment of the spirit that caused the illness (KII, Gbo) positioning traditional care not as a competitor to be displaced but as a culturally irreplaceable service. Table 5 maps these themes to the HBM–SEM framework.

**Table 5.**
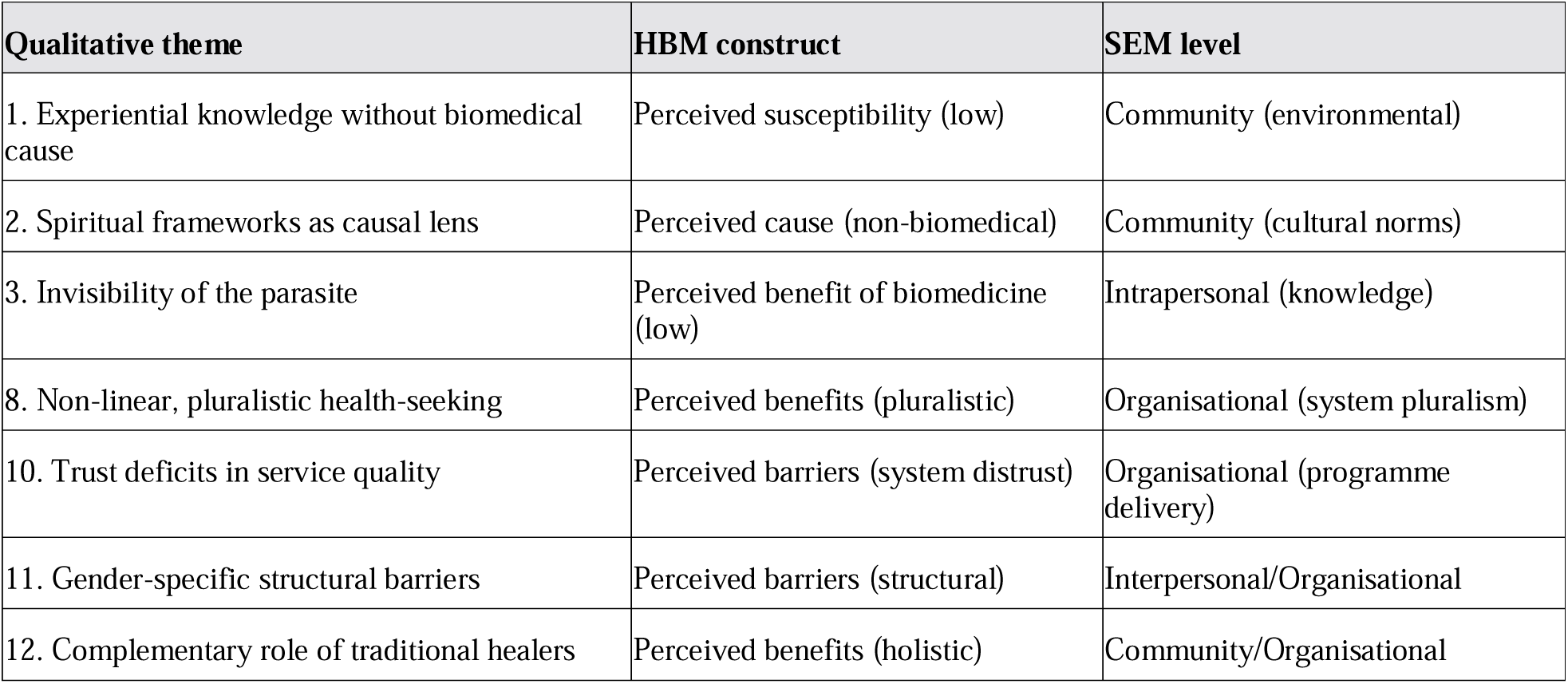
Integration of qualitative themes with the HBM–SEM framework.

| Qualitative theme | HBM construct | SEM level |
| --- | --- | --- |
| 1. Experiential knowledge without biomedical cause | Perceived susceptibility (low) | Community (environmental) |
| 2. Spiritual frameworks as causal lens | Perceived cause (non-biomedical) | Community (cultural norms) |
| 3. Invisibility of the parasite | Perceived benefit of biomedicine (low) | Intrapersonal (knowledge) |
| 8. Non-linear, pluralistic health-seeking | Perceived benefits (pluralistic) | Organisational (system pluralism) |
| 10. Trust deficits in service quality | Perceived barriers (system distrust) | Organisational (programme delivery) |
| 11. Gender-specific structural barriers | Perceived barriers (structural) | Interpersonal/Organisational |
| 12. Complementary role of traditional healers | Perceived benefits (holistic) | Community/Organisational |

## Discussion

In a large, representative sample drawn from five endemic chiefdoms of Bo District, biomedical knowledge of onchocerciasis was partial and uniformly so, non-biomedical aetiologies were dominant, and the traditional sector was the embedded first line of care. Crucially, neither perception nor the traditional-first pathway was patterned by chiefdom, sex or education in any programmatically useful way: the flagship model predicting traditional-first treatment was statistically null (McFadden R² = .005; AUC = 0.544). These are not failures of analysis but findings in their own right they show that the knowledge gap and the traditional-first pathway are generalised cultural defaults rather than the property of identifiable risk groups, and they explain why generic, demographically targeted health education has limited traction.

The partial-perception pattern symptoms recognised, parasitic cause misattributed mirrors knowledge attitude practice findings across endemic Africa [10,11,22](Okyere et al., 2024), but our data add the explanatory mechanism. The qualitative phase locates the deficit not in ignorance but in the literal invisibility of *O. volvulus* and in a culturally coherent attribution of disease to water spirits, curses and moral failing. Where the perceived benefit of ivermectin is filtered through a cosmology in which the true cause is spiritual, declining or deprioritising MDA is internally rational [12,13]. This reframes non-participation: it is not irrational non-compliance but a predictable response to a mismatch between the aetiological model promoted by CDTI and the one held by the community [9,12].

The prominence of the traditional-first pathway (55.3%) and the near-universal presence of healers (56.9%) confirm that care in Bo District is genuinely pluralistic, with biomedical and traditional modalities used concurrently rather than as alternatives [14,15]. The directional effect of distrust of modern medicine (OR = 1.33), set against a null demographic model, points to belief and trust not social position as the modifiable levers. Healers were described as offering meaning-making and psychosocial reintegration that the biomedical system does not, a complementary rather than competitive role [15,23]. Engaging healers as partners in referral and aetiology-aligned messaging is therefore more promising than attempting to displace them.

The structural and gendered barriers we document distributor trust deficits, the clash between morning distribution and women’s domestic labour, and inconsistent pregnancy messaging are consistent with the wider NTD literature on gendered access [16,24] and identify concrete, testable programmatic adjustments. Three follow: first, aetiology-aligned health communication co-designed with communities that engages, rather than dismisses, spiritual explanations and is delivered through trusted local channels; second, formal engagement of traditional healers in sensitisation and referral; and third, gender-sensitive MDA delivery, including afternoon distribution sessions staffed by female distributors and consistent, accurate messaging on ivermectin in pregnancy. Distributor accountability mechanisms — community scorecards and simple complaint channels directly address the trust deficits that the data identify as a binding constraint.

This study has limitations. The cross-sectional design precludes causal inference and cannot capture change over time. Self-reported knowledge and behaviour are subject to social-desirability and recall bias, partially mitigated by anonymity assurances, neutral consent framing and probing for negative experiences. Findings from rural endemic communities may not generalise to urban Bo City or non-endemic districts, and the post-conflict Sierra Leonean context may shape institutional trust in ways not directly transferable to other settings. The weak predictive performance of the flagship model, while substantively informative, also means that important determinants of treatment choice lie outside the individual-level variables measured here. Its strengths are a large, adequately powered probability sample, a pre-specified composite-index strategy, and the explanatory leverage of an integrated mixed-methods design.

In conclusion, the persistence of onchocerciasis in Bo District despite two decades of MDA is sustained not by demographic pockets of ignorance but by a community-wide configuration of partial biomedical knowledge, dominant spiritual aetiologies and an embedded traditional-first treatment pathway that drug distribution alone cannot dislodge. Closing the residual transmission gap will require elimination programmes to move beyond drug delivery to engage local explanatory models, partner with traditional healers, and dismantle the structural and gendered barriers that keep modern treatment out of reach. Addressing the social and cognitive determinants of treatment-seeking is not adjunct to elimination it is central to it.

## Acknowledgements

The authors thank the study participants, the Bo District Health Management Team, the chiefdom and village authorities, and the field research team for their support and cooperation.

## Data availability

The de-identified datasets supporting the conclusions of this article are available from the corresponding author on reasonable request and will be deposited in a public repository upon acceptance. Qualitative transcripts cannot be shared in full to protect participant confidentiality but are available in de-identified, excerpted form on reasonable request.

## Author contributions

**Conceptualization:** Augustine Bob Moseray, Alhaji Brima Gogra, Rashid Ansumana. Data curation: Augustine Bob Moseray. Formal analysis: Augustine Bob Moseray. Investigation: Augustine Bob Moseray. Methodology: Augustine Bob Moseray, Alhaji Brima Gogra, Rashid Ansumana. Supervision: Alhaji Brima Gogra, Rashid Ansumana. Writing original draft: Augustine Bob Moseray. Writing review & editing: Augustine Bob Moseray, Alhaji Brima Gogra, Rashid Ansumana.

